# Successive pandemic waves with more virulent strains, and the effects of vaccination for SARS-CoV-2

**DOI:** 10.1101/2021.09.21.21263901

**Authors:** Alcides Castro-e-Silva, Américo T Bernardes, Eduardo Augusto Gonçalves Barbosa, Wesley Dáttilo, Sérvio P. Ribeiro

**Affiliations:** Laboratório Ciência da Complexidade, Departamento de Física-ICEB, Universidade Federal de Ouro Preto, 35400-000, Ouro Preto/MG, Brazil; Departamento de Física-ICEB, Universidade Federal de Ouro Preto, 35400-000, Ouro Preto/MG, Brazil; Programa de Pós-Graduação em Modelagem Matemática e Computacional, Centro Federal de Educação Tecnológica de Minas Gerais – CEFET-MG. Av. Amazonas 7675, 30510-000,Belo Horizonte/MG, Brazil; Red de Ecoetología, Instituto de Ecología AC, Carretera Antigua a Coatepec, 351, El Haya, 91070, Xalapa, Veracruz, Mexico; Laboratory of Ecology of Diseases and Forests, Departamento de Biodiversidade, Evolução e Meio Ambiente-ICEB, Universidade Federal de Ouro Preto, 35400-000, Ouro Preto/MG, Brazil

**Author notes:** Email addresses:* (Alcides Castro-e-Silva), (Américo T Bernardes), (Eduardo Augusto Gonçalves Barbosa), (Wesley Dáttilo), (Sérvio P. Ribeiro).

**Keywords:** COVID-19, Pandemic, Vaccination, ABM-SIR model

## Abstract

Hundred years after the flu pandemic of 1918, the world faces an outbreak of a new severe acute respiratory syndrome, caused by a novel coronavirus. With a high transmissibility, the pandemic spreads worldwide, creating a scenario of devastation in many countries. By the middle of 2021, about 3% of the world population has been infected and more than 4 million people have died. Different from the *H*_1_*N*_1_ pandemic, which had a deadly wave and cessed, the new disease is maintained by successive waves, mainly produced by new virus variants, and the small number of vaccinated people. In the present work, we create a version of the SIR model with spatial localization of persons, their movements, and taking into account social isolation probabilities. We discuss the effects of virus variants, and the role of vaccination rate in the pandemic dynamics. We show that, unless a global vaccination is implemented, we will have continuous waves of infections.

**Highlights:**

- The COVID-19 has infected more than 200 millions and has killed more than 4 million persons.
- WHO has not been successful in defining a global vaccination policy.
- Many epidemic scenarios arise when different countries apply different vaccination strategies.
- Present model can show some insights on how vaccination programs can be managed.

## 1. Introduction

At the end of 2019 the world witnessed news about a novel disease that started in Wuhan, China. This illness, called COVID-19, is caused by a SARS class virus named SARS-CoV-2. Due to its high transmission capability, the disease rapidly reached all countries in the globe, mainly through airports networks [1, 2, 3], and, on March 11*th* 2020, WHO declared it as a pandemic. As of September 2021, the Johns Hopkins COVID-19 dashboard [4] showed more than 200 million cases and more than 4 million deaths globally. Such a number shows how fast the SARS-CoV-2 can spread through an entire population if no coordinated actions to prevent and reduce infections are not in place.

Any responsible sanitary policy adopted to slow down the progress of COVID-19 pandemic should get use of the following three strategies: a) social distancing: the obvious way to reduce susceptible-infected interaction and subsequent contagion, b) mask wearing and hygiene: once it is know that the transmission is mainly through respiratory droplets of infected patients and contact with surfaces infected by aerosol and, lately c) vaccines: a correct vaccination program can decrease the intensity of the disease symptoms among those infected but vaccinated, reducing the public health collapse risk, and the mortality rates, as susceptible but vaccinated people became asymptomatic. Still, the virus will circulate and the lack of a proper vaccination will create outbreaks due to contact between an increasing number of “asymptomatic” with vulnerable susceptible people. As [5] demonstrated, the existence of transient collective immunity may prolong an epidemic, and a bad vaccine scheme may exacerbate this pattern.

For the specific case of COVID-19 vaccination, one of the subjects of the present work, there are many factors that must be taken into account for a suitable immunization policy. Among all of them, this work will focus mainly on two aspects: how the virus is evolving into new variants and reinfection.

Every time SARS-CoV-2 infects a susceptible person, it starts to make copies of itself replicating its RNA [6, 7]. These changes or mutations in RNA can lead to different scenarios: it can be an evolutionary dead end and kill the virus, it can be an irrelevant and not noticeable change or it can bring some advantages, for example, better dealing with the immune system or better reaching the cells. Even more rarely, whole clusters of mutations can be acquired by the virus during a single infection. When a virus with single or cluster mutation gets spread through populations they are named “Variants of Concern” or VOC. According to CDC [8], a variant becomes a VOC when *there is evidence of an increase in transmissibility, or in lethality and severity of the disease, significant reduction in neutralization by antibodies generated during previous infection or vaccination, reduced effectiveness of treatments or vaccines, or diagnostic detection failures*. Although there are thousands of different genetic lineages of SARS-CoV-2, there are few Variants of Concern. Both the variant *β* (former B1.351), which was first detected in South Africa, and variant *γ* (former P.1) that was first seen in Brazil and Japan, contain mutations that appear to weaken the ability of antibodies to neutralise the virus by binding to it, which would normally prevent it from infecting cells. Variant *α*, (former B1.1.7), detected in UK and reported in 93 other countries and the variant *δ*, (former B.1.617.2), from India, shows less of an ability to escape from antibodies, but they have gained mutations that allows them to spread much faster than the original version of the virus.

Apart from VOC, there are still the “Variants of Interest” or VOI (variants *ϵ, η, ι, κ* and *ζ*) and “Variants of High Consequence” (there are no SARS-CoV-2 variants that rise to the level of high consequence until now). The CDC definition of a VOI is: *a variant with specific genetic markers that have been associated with changes to receptor binding, reduced neutralization by antibodies generated against previous infection or vaccination, reduced efficacy of treatments, potential diagnostic impact, or predicted increase in transmissibility or disease severity*. On the other hand, a variant has high consequences when it *has clear evidence that prevention measures or medical countermeasures (MCMs) have significantly reduced effectiveness relative to previously circulating variants*.

The second factor that can impact an immunization policy is the reinfection caused by the loss of immunity. Some works have shown that immunity with greater memory is acquired by infected people who developed severe symptoms, recovered and were vaccinated with at least one dose. However, even in these cases, immunity is not permanent, requiring a new vaccine [9, 10]. The reinfection was dimensioned as rare just before the spreading of the new variants of concern in early 2021 [11]. Still, not necessarily the loss of immunity in front of a variant of concern is a severe and frequent problem yet. However, risks may rise if the pandemic is not controlled and the virus is left to evolve freely [12, 13, 14]. Reduced neutralization of the Delta variant in comparison to the ancestral Wuhan-related strain was already observed [15], and a complex relation between different variants is also possible. For instance, it was also found that people infected with Beta variant are more susceptible to reinfection by Delta variant [15]. Hence, there is room for the evolution of new variants that could escape the vaccination more aggressively, especially if the vaccination scheme continues to follow a heterogeneous pattern, leaving the most vulnerable populations exposed for longer.

In this work, we developed an epidemiological model where events such as the appearance of new variants and reinfection are taken into account. Our results point to an optimal vaccine frequency that should be conducted in a given epidemiological setting.

## 2. The Model

In the present paper, we simulate a version of the SIR model [16] in a city with a population *N* (*t*) that may vary over time. Many variants of SIR model were used to simulate different scenarios of SARS-CoV-2 pandemy [17, 18, 19, 20], however, in order to have a better understanding of how cities’ structures and citizen dynamics affect the spreading of a transmissible disease like SARS-Cov-2, we choose to develop an SIR-ABM (Agent Based Model) [21, 22, 23, 24] of a “city” where their citizens start to get infected.

The city represents a geographically limited region. Only the arrival and departure of visitors and the deaths of its inhabitants can change its population. As in the previous versions of the SIR model, a *S*_*i*_ variable defines the health state of each individual: susceptible, infected, recovered, or immunized. In this work, we have included a fourth state: dead. Factors such as age, sex, or race are not taken into account.

In the simulated city, the residents live in houses, and they can move to public establishments (as malls, stadiums, stores, restaurants, etc). In some simulations, people can move to the houses of other people. The day starts with all the residents in their homes, to which they will be linked throughout the simulation. That is, if he/she leaves for another home or any establishment, at the end of the day, he/she will return to his/her home. Each person has a probability of movement *P*_*mov*_, and this probability defines social isolation mobility.

There is no natural movement, no duration of the action. The agents disappear from a place and reappear in another. We considered that the time of permanence in public or private places is longer than the travel time. On the other hand, for big cities, one can suppose that the time he/she stays in a mode of public transport, if prolonged, may also be considered as staying in a small public place with the same infection conditions.

According to the latest demographic census [25], Brazilian households have an average of 2.9 residents. In our simulations, we used an average of ∼ 3.3. In some simulations, the houses have either 3 or 4 residents, randomly chosen. In other simulations we detail below, we used an occupation given by a Poisson distribution with an average of 3.3 inhabitants.

Every public area has a carrying capacity of *K*, so that Σ*K*_*i*_ = *N*_0_, that is, the city supports that all its residents leave at the same time. There are several public areas with different *K*_*i*_ = {10, 000, 1, 000, 100, 50}, so that the sum of the capacity of each area of the same type is equal to 0.25*N*_0_. The number of places of a specific kind *i* is the total number of individuals they together can support divided by its *K*_*i*_. So, the number of sites that support fewer people is higher than those which support more people: there are more small stores than big stadiums. When individuals can move to other people’s houses, we define a carrying capacity of 12 individuals.

The time scale of our simulations is a day. For each day, individuals are chosen at random, in the number of alive people on that day. A resident either can not be drawn or could be drawn more than once. After this, we test if he/she will move, with probability given by *P*_*mov*_. Each individual may visit other places a maximum of three times during the day.

If the person is going to move, the places to which he/she will go are chosen at random: houses, small shops, or large stores. Its maximum capacity gives the likelihood of going to a location: it is more likely to go to a large store than to someone else’s home. If the selected place has its capacity reached, a new site is drawn until the person moves, ensuring that whoever was chosen to move will make a move.

The person arriving at a new location may be in one of three states: susceptible, infected, or immunized. If the person is vulnerable, one verifies if there is someone infected in the place. If not, the person stays there until a new movement or until he/she returns home at the end of the day. However, if there is infected person(s) at that location, first we calculate the probability of contact of this just arrived with an infected person, given by

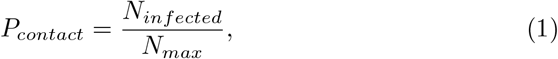

where *N*_*infected*_ is the total number of infected persons at that place (house or shop), and *N*_*max*_ is the maximum capacity of that place.

Two steps define the infection of a susceptible person: first, we calculate if he/she will contact a contaminated person. If so, one tests if he/she will have contagion. Contact does not imply contagion. The probability of contagion is *β*. In the case of COVID-19, *β* is estimated between [0.2, 0.3]. In the cases simulated in this work, we use a value of *β* = 0.2 for a “normal” variant and we assume more transmissible variants, with *β >* 0.2 [26].

If he/she acquires the illness, the probability of becoming infected with one of the variants present in that location is proportional to the frequency with which that variant is in the place, weighted by the transmissibility of the strain. The higher *n*_*i*_ ∗ *β*_*i*_, the higher the probability of being infected. *n*_*i*_ is the total number of individuals contaminated by the strain *β*_*i*_ at that location. The infected individual then becomes a potential transmitter of the disease.

If the individual who arrives at a place is infected or immunized *S*_*i*_ = 1 or 2, nothing happens to the others. The arrival of an infected person will create the conditions for those who arrive later to be infected.

At the end of the day, after performing some movements, all individuals return to their homes. Of course, if he/she has not moved, nothing is tested. However, for those who return to his/her home, the same set of procedures to check for infection is adopted: If the person returning home is susceptible, and someone infected is in the house, the same steps described above are carried out for contact and contagion.

There is an asymmetry in the model. If an infected person arrives at a place, we do not test if he/she will infect the susceptible ones already there. This kind of procedure corresponds to a sequential order in the contact/contagion process. It is understood that for huge populations and many days of simulation, this fact does not imply results different from those reported below.

The state of infection in an individual remains for a maximum of 1*/γ* = 14 days. It is assumed that the potential for infecting another person does not change during this period. It depends only on the *β* value, which does not change for a contaminated individual.

After seven days of infection, an individual has a probability of death of *P*_*death*_. If a more transmissible variant infects him/her, the likelihood of death increases by *δ*_*beta*_, changing *P*_*death*_ of that individual. This antagonism creates a tension between the strain’s transmissibility and the individual’s probability of death. That is, hosts with a more transmissible variant are more likely to die.

We have also simulated the case of incubation. In this case, the susceptible become infected after five days of incubation. During these five days, they can not infect other individuals.

After 14 days, an infected individual becomes an immunized or recovered one. He/She remains in this immunized state for *T* days when he/she returns to the condition of being susceptible. During this period, he/she can not be infected with any variant of the new coronavirus [27, 28].

Our model assumes that visitors arrive and leave the city every day. The number of visitors coming and leaving each day is around 1/1,000 of the total residents. These visitors have a probability of movement equal to 1. It means that a visitor will go to three places (houses, small shops, or large stores). The likelihood of a visitor being infected is 0.1%. In principle, if infected, he/she will have the virus variant with *β* = 0.2, but some can have a more transmissible variant. We have implemented a version in which visitors are 1% of the population. Of the visitors, 3% may have the new type of virus.

In this model, there are no births. There is no increase in the number of inhabitants, except visitors, but it represents a zero change in the population since they arrive and leave. The only factor that can change the resident population is the death caused by disease. We study the effects of a new variant of the virus, which enters via visitors, studying the competition dynamics between different strains. Besides, we also want to study the effects of vaccination, not considering other variables. A local variant is not supposed to mutate.

If that visitor is susceptible, he/she may become infected during the day and transmit the infection. But as he/she only spends a day in the city, he/she will not be subject to death, nor will he/she be able to recover. He/She will also not be vaccinated.

In our simulations, vaccination starts from the 300^*th*^ day. This date roughly corresponds to the beginning of vaccination campaigns in several countries: between December / 2020 and January / 2021, considering the detection of the pandemic as time zero (around February or March 2020). A *P*_*vac*_ percentage of the population is vaccinated each day. When vaccinated, an individual acquires immunity. There is no need to use two doses or intervals between doses of the vaccine to gain immunity. In this simplified version of the model, immunity is acquired at the time of vaccination. An immunized individual will not be infected by any other variant of Sars-Cov-2 (referencias). After *T* days, the immunized individual returns to the condition of being susceptible.

We will show the results with two versions of the model: in the first one, we simulate a city with houses, small and big stores, where the visitors may carry just one virus variant with higher transmissibility. Residents can move to other residents’ houses; in the second version, the towns have homes and just one type of store, and the visitors carry many virus variants (described below).

In the first version, we studied the effects of vaccination, and it was not in the second version. The central aspect of the first version is to look at the spread of a higher transmissibility strain with vaccination; the second version aims to study the competition between different strains. In both cases, we consider that variants with different transmissibility also have different lethality.

## 3. Results

### 3.1. Model with two strains

The first set of results have been obtained in simulations of cities with two populations: *N* ∼ 875 thousand residents, and another with *N* ∼ 8.75 million residents. In the first case, they live in 250 thousand households. When allocating people, it is drawn whether the house will have 3 or 4 residents, giving 3.5 residents on average per household. These residents can move to 20,000 stores with a maximum capacity of 50 people or 2,000 establishments which support 500 people. Each kind of place (houses, small or big shops) can admit all people’s movements, which means that there is space in any space for moves of all people if it would be the case.

Each day, people can leave the house with a movement probability of *P*_*mov*_. In the simulations described below, values of *P*_*mov*_ = 0.6, 0.5, *or*0.3 were adopted, meaning an average isolation of 40 %, 50 % and 70 %. In the case of Brazil, the average isolation, measured while individuals stay in their homes for a day, has fluctuated between 30 % to 40 %, and on weekends it can reach 50%. It increased a little in February and March 2021 due to the imposition of new measures of restriction and operation of the establishments. At the beginning of the pandemic, in March 2020, there were higher values in Brazil, in the range of 60 %.

For each time step, individuals are chosen at random, as described above. One verifies if he/she will leave the house. If so, we draw the total number of moves he/she will make: 1 up to 3. Then, we randomly choose the places for visitation: other houses or small or large stores. He/she leaves the house and goes to each establishment. Each person executes his/her movements, and after completing them, stays at the last site until he/she returns home at the end of the day.

In all the simulations discussed below, a portion 10% of the population has a probability of movement of *P*_*mov*_ = 0.1, representing the people who are most at home. As described above, visitors come and go. We have assumed a proportion of visitors of 0.1 % of the total initial population per day in the results we show below. These people who arrive and leave remain only one day in the city. A ratio of 1/1,000 may be contaminated. The contamination strain has *β* = 0.2. A portion of 1*/*90 of the contaminated may have a more aggressive variant, with *β* = 0.25. Visitors have a probability of movement equal to 1.0 and will always visit 3 locations (drawn at random as described above).

Each individual *i* is assigned a variable *S*_*i*_ that can be in four states: susceptible (0); infected (1); recovered or immunized (2); and dead (3). People start the simulation as susceptible: all with *S*_*i*_ = 0. A single person, among the residents, at *t* = 0 is infected with the less aggressive variant *β* = 0.2. The dynamics is given by the process shown in Figure 1.

**Figure 1:**
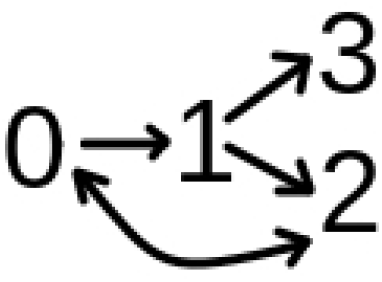
Schematic representation of transitions between states in our model. Probabilities are discussed in the text

The figure represents the possible changes of state. A susceptible individual *S* = 0 can become infected *S* = 1 with probability given *P*_*contact*_ × *β*_*i*_. *β*_*i*_ is the transmission rate of an individual which contaminates that susceptible, as described above. An infected individual *S* = 1 can die *S* = 3 if his/her time of infection is greater than 7 days and with probability of death given by *P*_*death*_. In case of being contaminated with the most aggressive variant *β* + *δ*_*β*_, the value of *P*_*death*_ is increased by *δ*_*death*_. After 1*/γ* days, the infected individual *S* = 1 becomes recovered or immunized *S* = 2. After *T*, the immunized individual *S* = 2 returns to the condition of susceptible *S* = 0. With probability *P*_*vac*_, a susceptible or infected individual, either *S* = 0 or *S* = 1, can be immunized *S* = 2, simulating the fact of having been vaccinated. Immunization protects an individual for *T* days. We show simulations with *T* = 120, or 180 days.

In the results shown below *P*_*contact*_ is given by equation (1), *β* = 0.2, *δ*_*β*_ = 0.05, *P*_*death*_ = 0.01, *δ*_*death*_ = 0.004, 1*/ gamma* = 14, and *T* = 120 days.

On the simulation’s 300th day, a vaccination campaign may start. We performed simulations with two vaccination rates. Some simulations used a vaccination probability of *P*_*vac*_ = 1*/*200, representing the typical value of vaccination campaigns in Brazil when close to 1 million people are vaccinated per day. Brazil has a public health system formed by about 40 thousand health centers belonging to three levels of administration but forming a cooperative network. This system has vast experience on vaccination campaigns and can easily reach the rate of 1 million vaccinations per day, roughly 1/200. The second value we have tested is a rate of 1/1,000, which represents values that we have observed in vaccination against COVID-19 in the first two months of vaccination, close to 200 to 300 thousand people per day. From January 23, 2021, to March 29, 2021, 14 million people received the first dose of one of the two available vaccines. In Brazil, the vaccines used are those that require two doses in their application. It increased in April but oscillated since Brazil has no plan of vaccine acquisition. In the USA, the vaccination rate reached 0.006 of the whole population in March. By the beginning of March, Israel had already vaccinated practically its entire population of around 9 million people.

Figure 2 represents the evolution of a population with 40% of social isolation: *P*_*mov*_ = 0.6, without vaccination. The values represent the densities relative to the initial number of residents, in this case, 874,975 people. The legend identifies each curve representing a specific *S* state. There is an oscillating evolution of the numbers caused by the permanent entry of infected individuals: the visitors. The curve called *β*_*av*_ represents the average value of *β* taken among all individuals with *S* = 1. It is observed that in the peaks of infection, the most aggressive variant tends to dominate. The decrease in the infected population is typical of the SIR model, but the permanence of the most aggressive variant is an important characteristic. The number of individuals who die is in the order of 10 % of the original population. Certainly, it is a very high number when compared to real data. This model and simulations don’t intend to project expected values, but they represent the dynamics of disseminating the most aggressive variant qualitatively.

**Figure 2:**
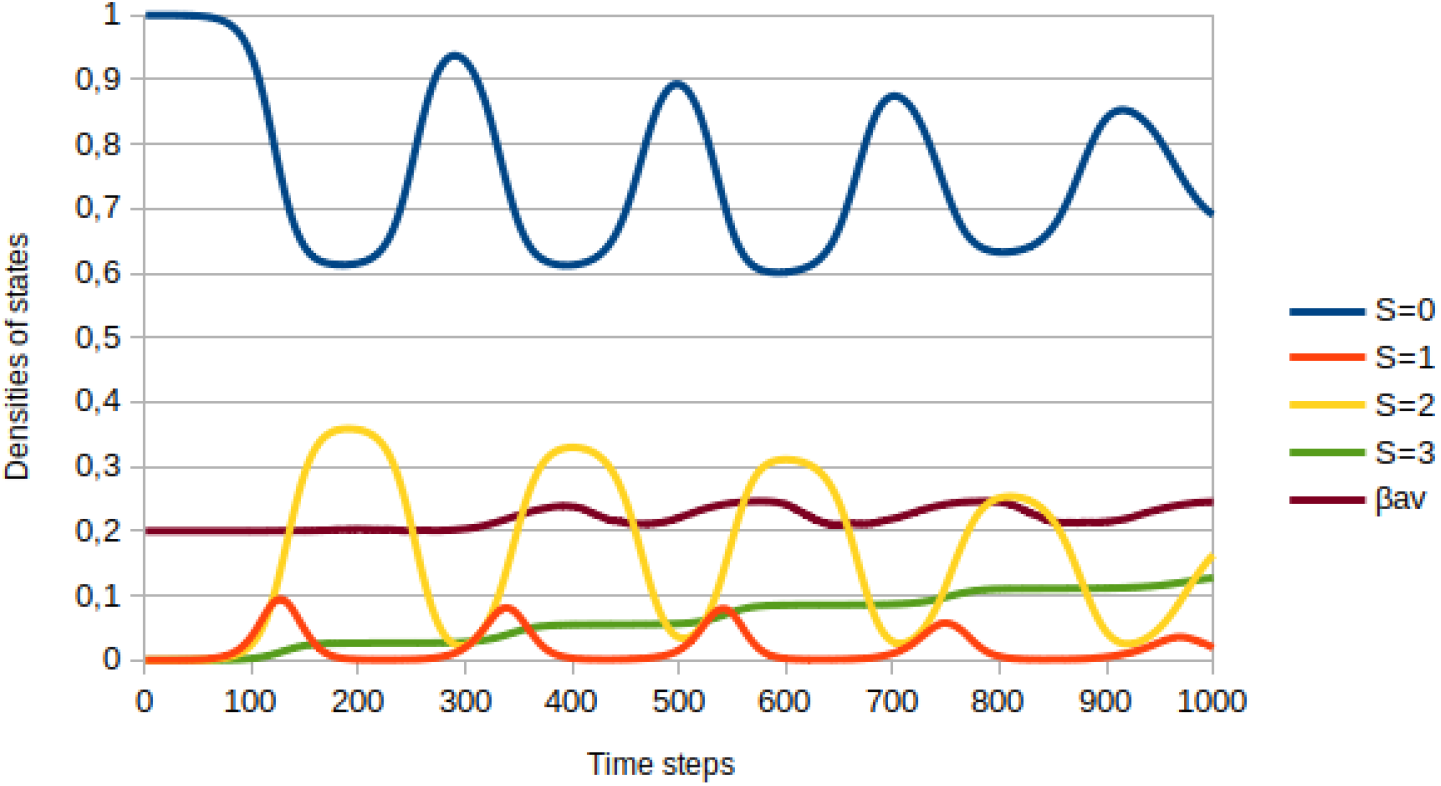
Evolution of densities of states. The legend defines the meaning of colors. Initial population is ∼ 8.75 × 10^5^

Figure 3 shows the percentage of people dying each day (blue line) and among these the percentage of people who died by the most aggressive variant (red line). The results have been in the simulation shown above. One observes that the number of people who die each day increases in the periods of infection. However, the quantity of those who die from the new variant rises more rapidly, becoming ∼ 50% at the end of the simulation.

**Figure 3:**
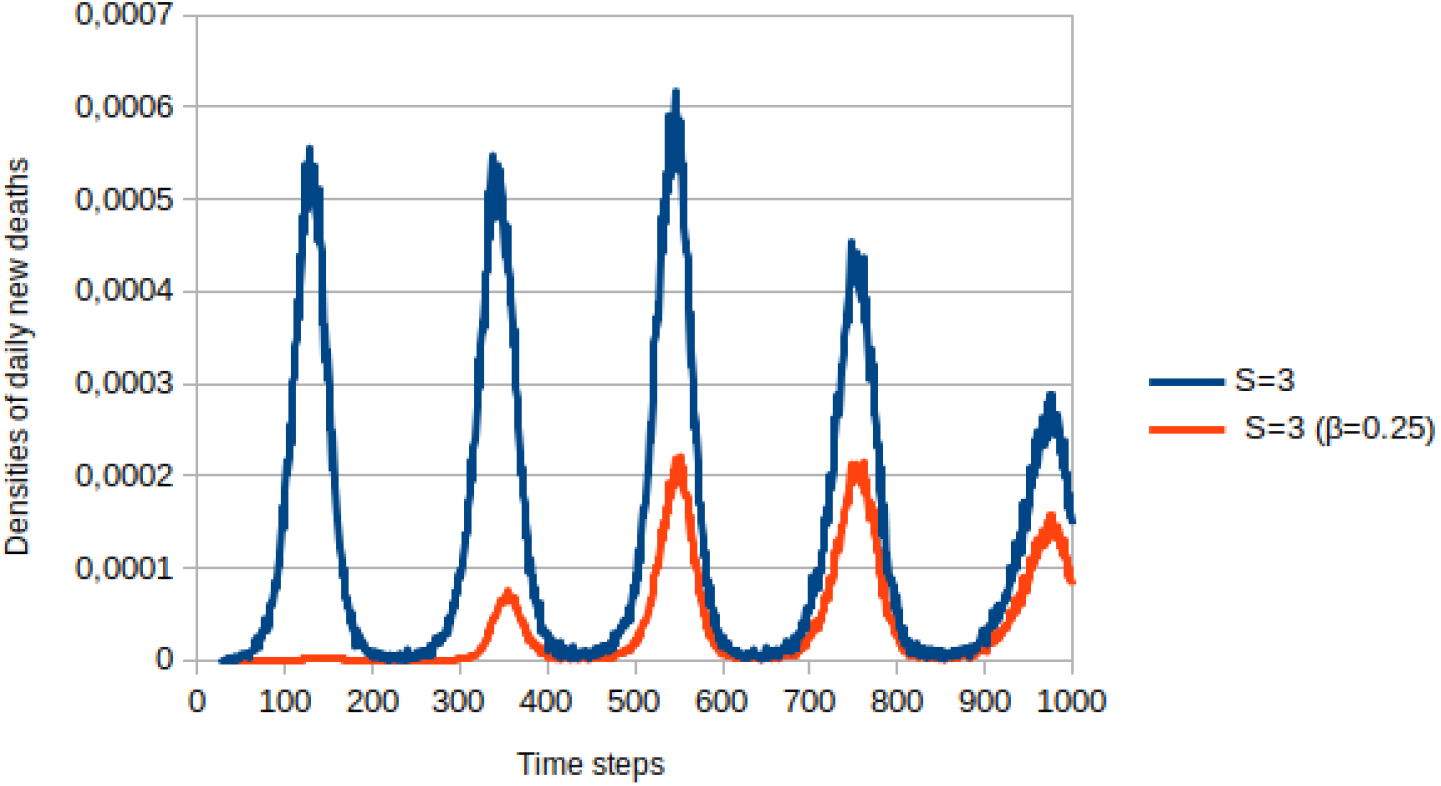
Density of daily new dead individuals - blue line, proportion of individuals who died by the most lethal variant - red line

Change in this kind of dramatic evolution is only achieved when we have about 70 % of social isolation. For this result, we do not have the vaccination. Figure 4 shows a simulation obtained with the previous parameters, but now with *P*_*mov*_ = 0.3. In this case, the susceptible population remains basically between 0.95 and 1, the density to the original number of residents. Infected people cannot contaminate significant fractions of the population, and the infection stops. This social isolation of 70 % of isolation is discussed and defended in many papers and speeches as the necessary value to stop the pandemic since there is no previous or preventive medication. But even in this case, the most transmissible strain dominates, as one can observe from the average value of *β*.

**Figure 4:**
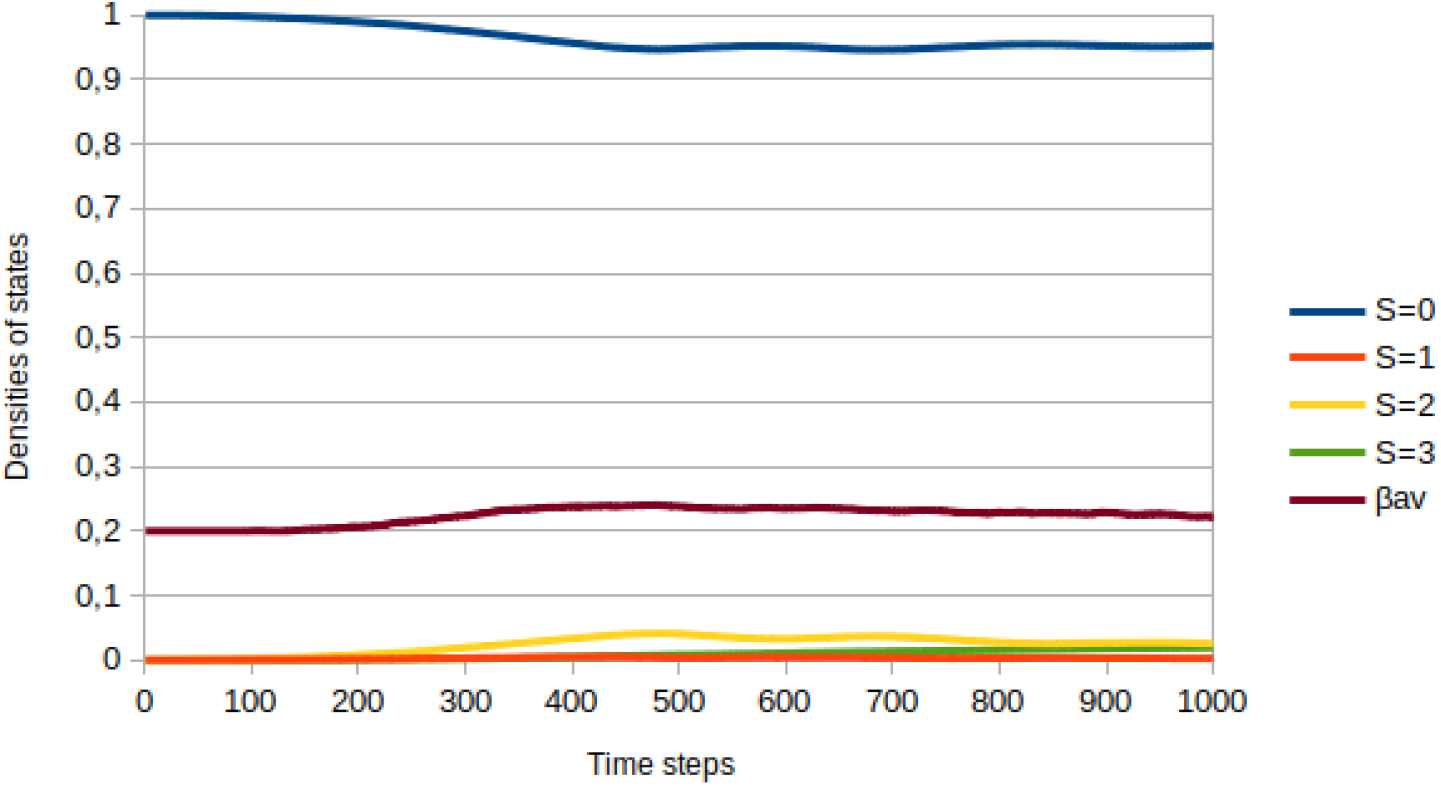
Evolution of densities of states for social isolation of 70%, *P*_*mov*_ = 0.3. The legend defines the meaning of colors. Initial population is ∼ 8.75 × 10^5^

Brazil has never achieved these higher values of social isolation. Besides the economic situation of the most vulnerable part of the population, there is robust dissemination of fake news from government authorities or even from digital influencers, defending the existence of early treatment, combating basic sanitary measures, such as using masks. Recently, a work discussed the effect of opinions and information on pandemic dissemination (quote from recent Physica A). Opinions, mainly from political leaders, have a significant influence on the dynamics of the pandemic.

After showing the results obtained without vaccinations, we present the studies when there is vaccination, with two vaccination rates: 1/200 of the population, meaning a more significant portion of the people. The second value used in the simulation for vaccination rate is 1/1,000 of the population per day.

Figure 5 shows the result obtained with vaccination at the rate of 0.001, with a probability of movement of 60 % corresponding to the social isolation of 40 %, peak value currently in Brazil. The numbers of infected and the progression of the pandemic do not stop. The numbers of deaths and the spread of the most aggressive variant follow the patterns observed previously. It happens because the rate of vaccination, that is, the transition from susceptible and infected to immunized, is low compared to the rate of change from vaccinated to susceptible. Note that the time for this transition is *T* = 120 days, meaning four months in our time scale.

**Figure 5:**
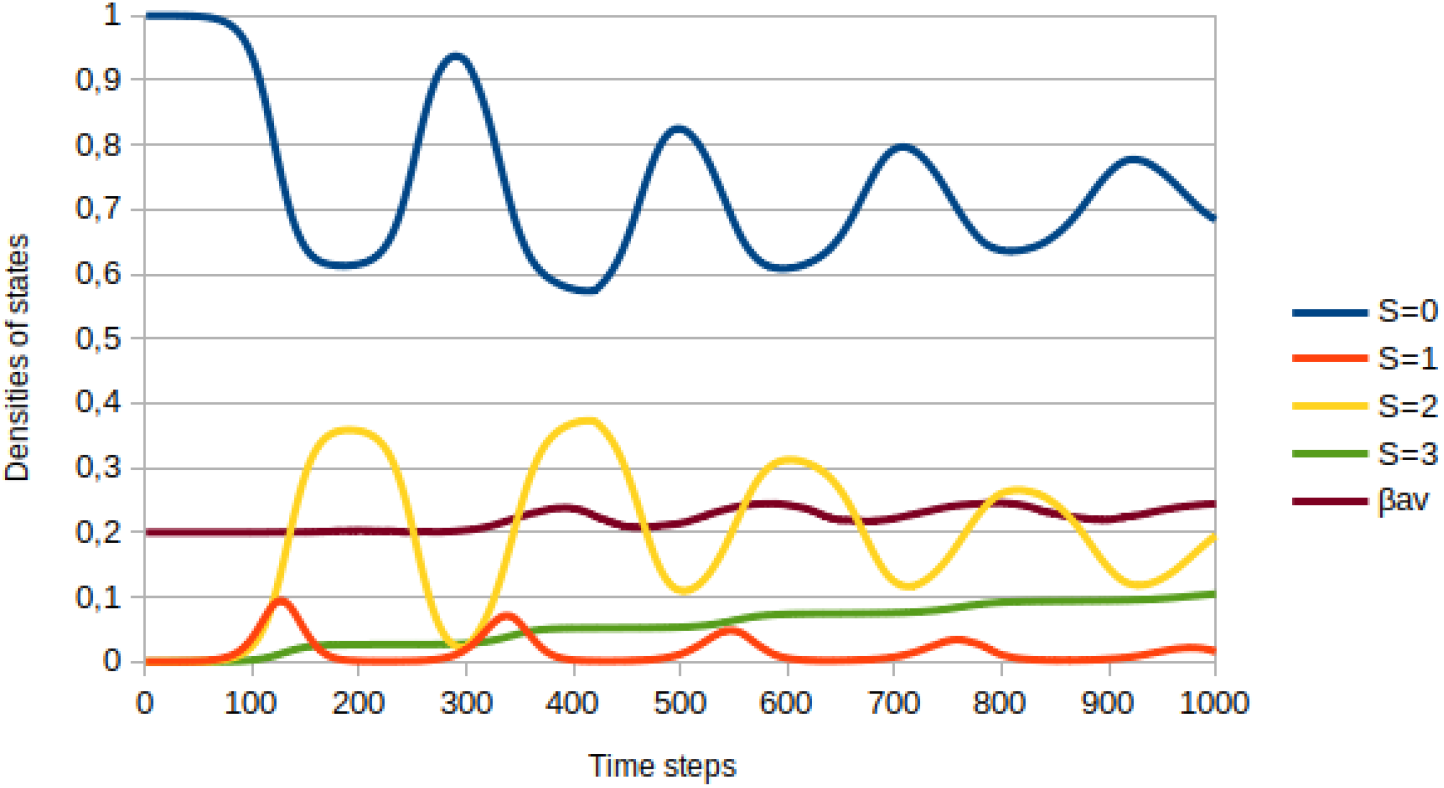
Evolution of densities of states for *P*_*mov*_ = 0.6, and vaccination rate 0.001. The legend defines the meaning of colors. Initial population is ∼ 8.75 × 10^5^

When the immunization rate assumes the value of 1/200, the picture changes radically, as shown in Figure 6. Vaccination started on day 300 of the simulation. One observes that there is still the second wave of infections because the virus is widespread in the population. But the number of immunized people increases rapidly, which is a blocking factor for the spread of the pandemic. One also observes that the most aggressive variant is contained, as the value of *β*_*av*_ is closer to 0.2. It is important to remember that the rate of contaminated visitors is the same in the three cases discussed so far. The number of deaths also stabilizes with the blockade caused by vaccination.

**Figure 6:**
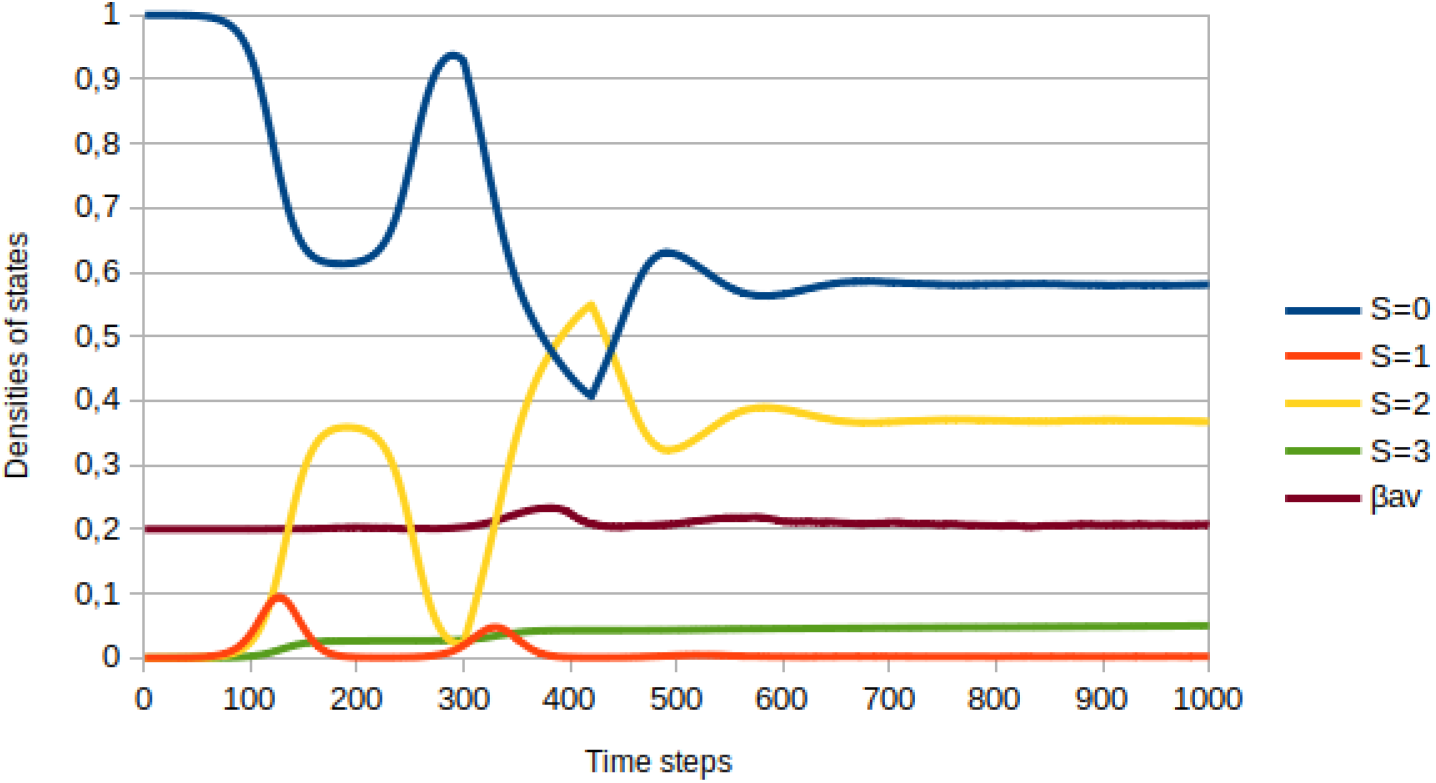
Evolution of densities of states for *P*_*mov*_ = 0.6, and vaccination rate 0.005. The legend defines the meaning of colors. Initial population is ∼ 8.75 × 10^5^

The following Figure 7 reproduces the relationship between daily deaths and the presence of the most aggressive variant. This variant is still present in the second wave but on a smaller percentage than in Figure 3. Later, this variant becomes marginal. It is necessary to remember that this most aggressive variant enters the population from the visitors, and therefore there will always be, in this model, the presence of this variant.

**Figure 7:**
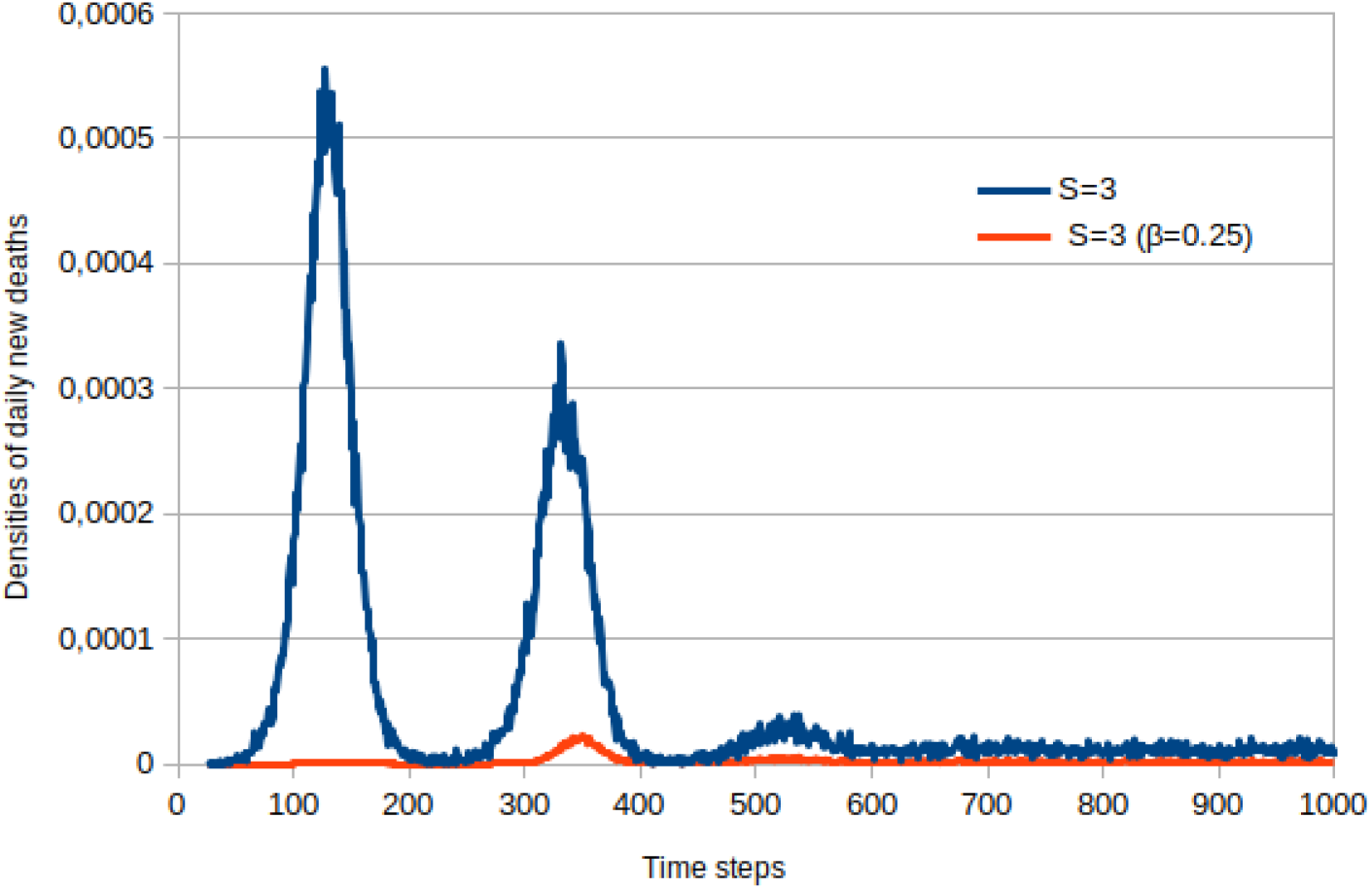
Number of dead individuals per day - blue line, number of individuals killed by the most lethal variant - red line. The figure shows results for *P*_*mov*_ = 0.6, and vaccination rate 0.005.

Figure 8 summarizes this discussion, showing the number of infected people according to isolation and vaccination rates. The first figure (A) shows the case where there is no vaccination. One observes that only with an isolation rate of 70% the pandemic is stopped. In the second case (B), when the vaccination rate is 1/1000, it is observed that there is no significant impact on infection dynamics. Only with a higher vaccination rate (C), as mentioned in the cases of Israel or the USA, can the pandemic be stopped, even with lower isolation rates.

**Figure 8:**
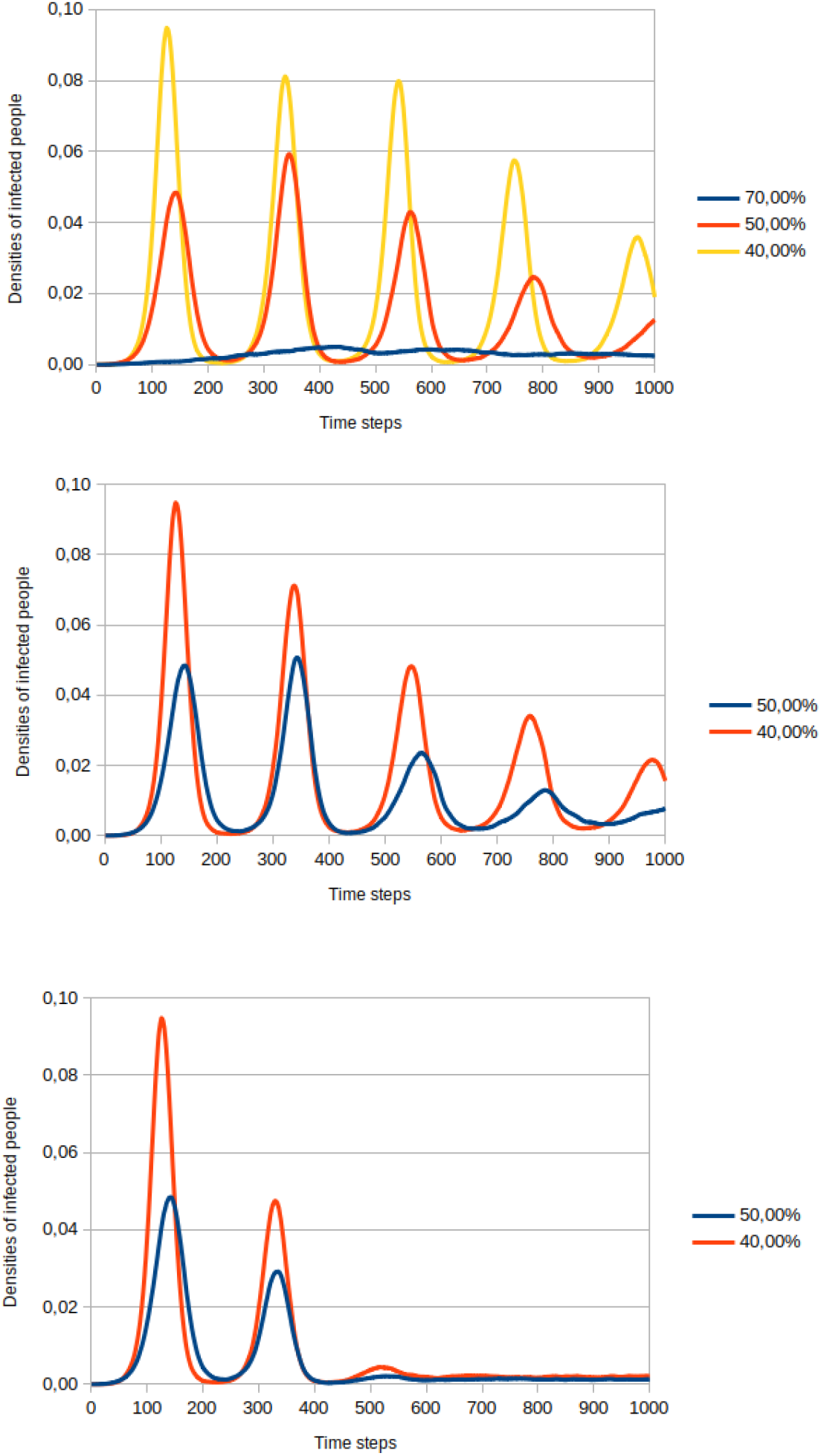
Comparisons between three groups of simulations. Only the density of infected people is shown. All the simulations refer to the initial population of ∼ 8.75 × 10^5^, and general parameters described in the text. Different probabilities of movement are defined in the legends. Part (A) represents simulations without vaccination; part (B) with vaccination rate of 0.001; and part (C) with vaccination rate of 0.005

To better understand the influence of the city size, the number of individuals, as well as the period of immunization after vaccination, we performed simulations with *N* 8.75 million individuals and *T* = 180 days, meaning that a recovered or immunized stays in the *S* = 2 state for six months.

All the parameters are the same as those used in the simulations described above: *P*_*mov*_ = 0.6, *β* = 0.2 for residents, number of visitors 1/1,000, the proportion of contaminated visitors 1/1,000 being those with the highest transmissibility rate 1/90. Vaccination, when it occurs, starts on the 300*th* day.

In Figure 9, the curves represent only infected (red line) and deaths (green line) densities. Part (A) represents the simulation without vaccination; part (B) represents vaccination starting on the 300*th* day, with a 1/1,000 of vaccination rate; and part (C) represents the evolution with a 1/200 vaccination rate. Qualitatively, the behavior is the same as discussed above for populations ten times smaller. The main aspect is that the change in the immunization period does not change the evolution of successive waves for the cases without vaccination or with a low vaccination rate.

**Figure 9:**
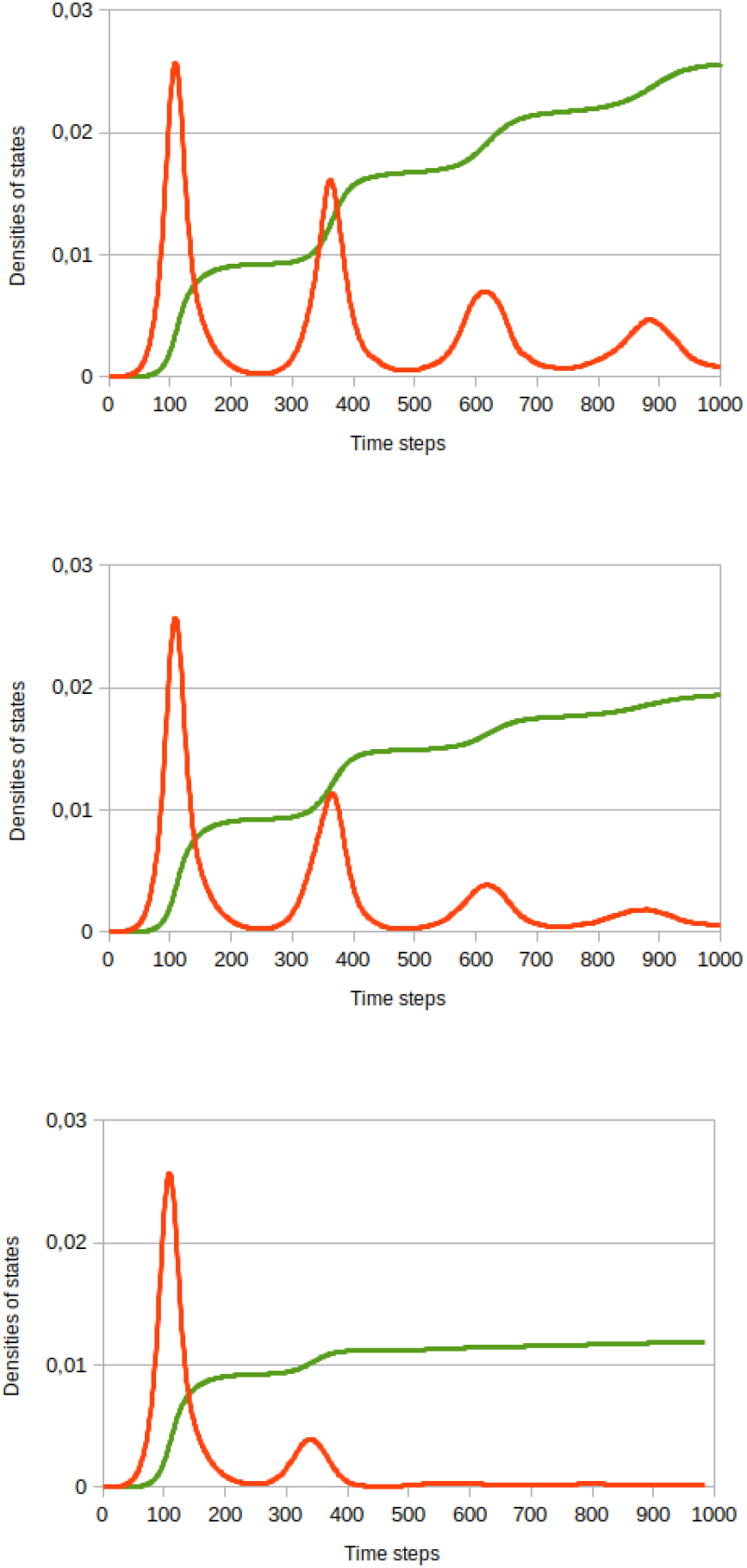
Comparisons between three groups of simulations for large population ∼ 8.75 × 10^6^, and immunization period *T* = 180 days. *P*_*mov*_ = 0.6 for all these simulations. Only the densities of infected people - red line, and death people - green line. The other parameters are described in the text. Part (A) represents simulations without vaccination; part (B) with vaccination rate of 0.001; and part (C) with vaccination rate of 0.005

### 3.2. Model with many strains

We have implemented a second version of the model where residents can move only to public places. We have considered a more significant number of visitors who can bring different variants of the virus. In this version, for each instant of time (day), residents leave their houses with a probability that defines social isolation. They stay at home or move to public areas, making 1 or 2 movements per day. We simulated the initial isolation of 45% as default. In this second model the flux of visitors is 1% of the total population, but infected travelers are 3% of the total number of visitors. All the other parameters are the same as the original model. The objective of this second model is to simulate how different strains compete among themselves in an epidemic scenario. This is done in the following way: the simulation starts with just one type of variant (called variant 1 on plots), after 5% of population gets infected, 4 other variants (2, 3, 4, 5) are introduced via infected visitors. These variants have different contagion and mortality probabilities as can be seen in table 1. It shows that, compared with the original strain, we have a combination of 70% and 50% more and less contagious and lethal variants respectively.

**Table 1:**
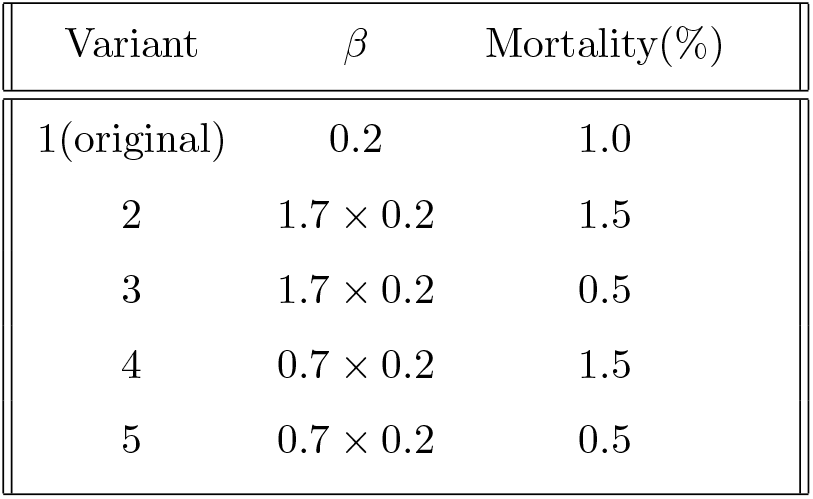
Contagion (*β*) and mortality of original and variant strains used in the second model.

The results are presented in figures 10 and 11.

**Figure 10:**
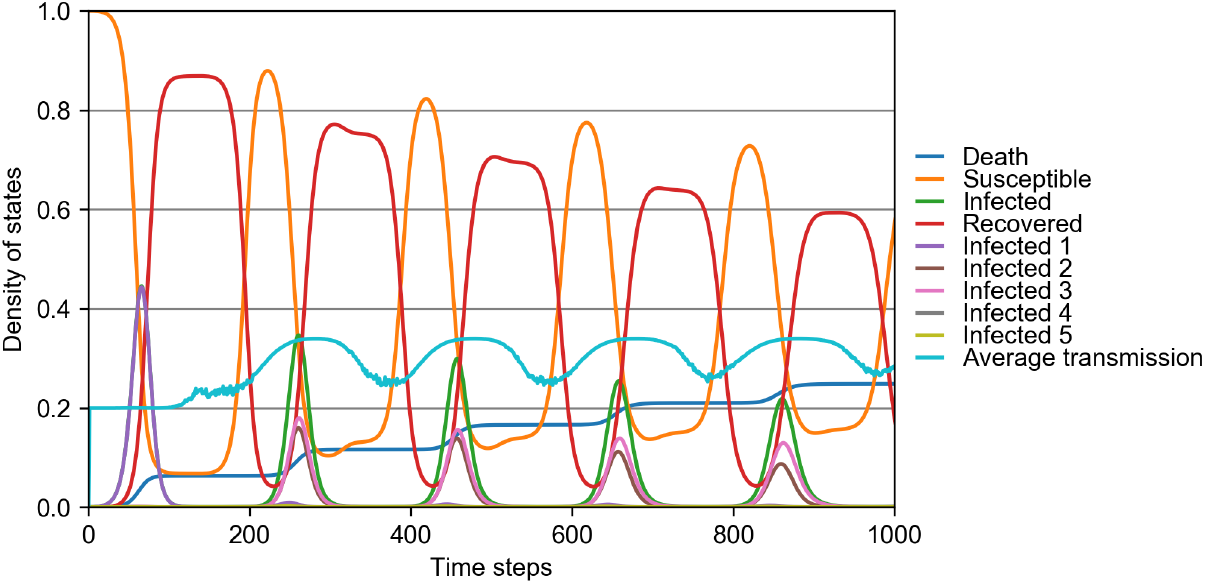
Evolution of densities of states for a population of 1M, with 1 variant entering via travelers. The social distancing is 45%. The values of *β* and mortality for infected can be seen in table 1. In this scenario, all recovered lost their immunity after 120 days.

**Figure 11:**
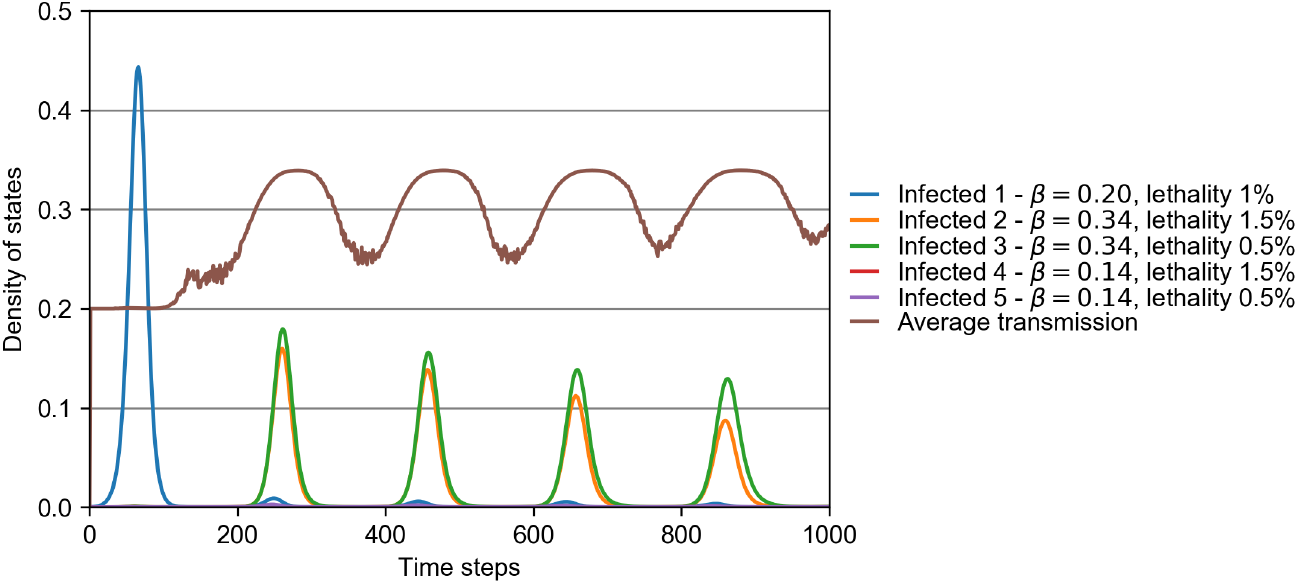
Presence of virus in population, same scenario of figure 10. This plot show how rapidly infection caused by variants 2 and 3 rapidly overcome the original one from the second wave on.

Figure 10 shows the density of states in a simulated city with a population of 1 million, with a 45% of social distancing, and a variant more contagious entering via travelers. The result shows the waves of infection caused mostly by loss of immunity after 120 days in a similar way to the results of the first model without vaccination. The competition among different variants can be better viewed in figure 11. This plot shows, for the same parameters of the previous plot, how the different strains are present in the population during time. It can be seen how the original strain causes a peak of ∼ 45% of people infected in a first wave around the day 80. However, once the four new variants are introduced in the system, the following waves show that the original variant are rapidly replaced by the two most transmissible ones (variants 2 and 3 of table 1). Since the second wave (starting between days 200 and 300) the simulation shows a dominance of variant 3 (more transmissible, less lethal), followed by variant 2 (more transmissible, more lethal) and finally a barely presence of original variant. This plot shows two important results: (A) the new variants with less transmissibility than the original strain (variants 4 and 5) are not able to invade the city while the more transmissible variants substituted the original one from second wave on and (B) interestingly, the lesser lethal variant overcame the more lethal one in all the waves, which is expected for a long-term evolution of an emergent disease if the lack of host illness favors transmission, i.e., loss of virulence [29, 30]. Due to the full shifting of the original strain by new variants, the *β*-infection rate was permanently higher after the second wave than it was in the beginning of the pandemics, as shown by average transmission on figure 11.

## 4. Conclusion

After some waves of infections, the world is facing a new challenge. Some countries, like the USA, have a significant part of the population which refuses to be vaccinated. This likely allows the spread of new variants and they now observe a fifth wave of infection, this time much more intense than the previous one, which occurred in the middle April 2021. Vaccination slowed down and this may cause the present situation. In Israel a new wave is observed. But since they have a high vaccination rate, the number of deaths is smaller than in the previous wave. Brazil presents a different situation. The country started vaccination with a low rate, due to the role of the federal government. However, there was a strong engagement of the population and, after public pressure from but also the action of political sectors, the rates of vaccination increased in the last months. The number of contaminated and dead people remained in a plateau during the first months of this year, but declined since the augment in the vaccination rate. Different scenarios can be observed in different countries. However, they are a product of the spread of new variants and the population vaccination.

In this paper, we simulated the competition between virus strains, the role of the vaccination rate, and social isolation, which is believed as one of the main aspects to detain the virus’s circulation.

As in many countries, like Brazil, it is very complicated to maintain social isolation for long periods. We observe stress in parts of populations children out of schools, increase in domestic violence and difficulties to obtain the basic income to sustain the families. Poverty increased in many countries. In the particular case of Brazil, social isolation never reached the adequate or recommended levels, which is responsible for the long plateau of infections and deaths.

We show, thus, that the only way to stop the virus circulation, or at least to diminish the contamination rates is increasing the vaccination rates. Low vaccination rates allow the circulation of the many variants and we observe a cyclic problem. The situation gains a much more dramatic feature with the existence of new variants with a higher transmissibility.

The many different variants will compete and those with higher transmissibility will win the competition, even with a higher lethality. This picture is observed where the *δ* variant appeared. This variant rapidly infests people, even those already vaccinated and can cause a higher increase in the total of contamination. This led us to the problem in the beginning of the pandemic, which was how to flatten the infection curve. Because the spread of new strains can overload the healthy systems.

The main purpose of this paper is to shed some light in the main aspects of the actual dynamics of the pandemic. It is clear that one needs a global governance to deal with this dynamic, because the total curve of the pandemic dynamics exhibits successive peaks with a distance of about 4 months between them. So, without global control, new variants continue to appear and the infection is partially controlled in one country or region, but increases in other parts, in a dramatic cyclic death wheel.

## Supporting information

Fortran 95 code for model #1

C++ code for model #2

## Data Availability

The code of model is included in manuscript files.

## Acknowledgments

This research did not receive any specific grant from funding agencies in the commercial, or non-profit sectors. SPR acknowledges grant from Conselho Nacional de Desenvolvimento Científico e Tecnológico - CNPq, through proccess 306572/2019-2. The authors declare that there are no conflicts of interest.

